# Benchmarking pathology foundation models for non-neoplastic pathology in the placenta

**DOI:** 10.1101/2025.03.19.25324282

**Authors:** Zehao Peng, Marina A. Ayad, Yaxing Jing, Teresa Chou, Lee A.D. Cooper, Jeffery A. Goldstein

## Abstract

Machine learning (ML) applications within diagnostic histopathology have been extremely successful. While many successful models have been built using general-purpose models trained largely on everyday objects, there is a recent trend toward pathology-specific foundation models, trained using histopathology images. Pathology foundation models show strong performance on cancer detection and subtyping, grading, and predicting molecular diagnoses. However, we have noticed lacunae in the testing of foundation models. Nearly all the benchmarks used to test them are focused on cancer. Neoplasia is an important pathologic mechanism and key concern in much of clinical pathology, but it represents one of many pathologic bases of disease. Non-neoplastic pathology dominates findings in the placenta, a critical organ in human development, as well as a specimen commonly encountered in clinical practice. Very little to none of the data used in training pathology foundation models is placenta. Thus, placental pathology is doubly out of distribution, representing a useful challenge for foundation models.

We developed benchmarks for estimation of gestational age, classifying normal tissue, identifying inflammation in the umbilical cord and membranes, and in classification of macroscopic lesions including villous infarction, intervillous thrombus, and perivillous fibrin deposition. We tested 5 pathology foundation models and 4 non-pathology models for each benchmark in tasks including zero-shot K-nearest neighbor classification and regression, content-based image retrieval, supervised regression, and whole-slide attention-based multiple instance learning. In each task, the best performing model was a pathology foundation model. However, the gap between pathology and non-pathology models was diminished in tasks related to inflammation or those in which a supervised task was performed using model embeddings. Performance was comparable among pathology foundation models. Among non-pathology models, ResNet consistently performed worse, while models from the present decade showed better performance. Future work could examine the impact of incorporating placental data into foundation model training.

## INTRODUCTION

Machine learning (ML) for pathology diagnosis has been remarkably successful, with FDA clearance of diagnostic models for prostate and breast cancer resulting in clinical impact.^1,2^ In the last 2-3 years, the field has moved away from using models pretrained on diverse, generally non-medical images to building pathology domain-specific foundation models, often based on vision transformer (ViT) architectures (**Table 1**).^1–9^ These models are trained on datasets of thousands to millions of slides. Each slide may have tens of thousands of 224×224 pixel patches. The amount of data dwarfs the 1.4 million images in ImageNet and even exceeds the 142 million images in the general purpose ViT foundation model, DINOv2.^10^ More complicated models, with larger numbers of parameters, can take full advantage of larger training datasets to maximize performance.^4,11–13^ The combination of large datasets and large models results in long, expensive training times. In exchange for this cost, foundation models show several advantages. Foundation models can be used as the basis for many diagnostic models, potentially simplifying training, and allowing for embed-once, infer many approaches in deployment. Fine-tuning pathology foundation models might be expected to yield higher performance than fine-tuning models pretrained on non-medical images. Finally, foundation models show the ability to perform zero-shot tasks – that is, they may have acceptable performance on new tasks without additional training.

**Table 1.** Selected foundation models: Pathology models are above the line. TCGA: The cancer genome atlas.

| MODEL NAME | Citation | Training data | Tissue sites | Model parameters | Data source |
| --- | --- | --- | --- | --- | --- |
| CONCH | Lu, 2024 | 1.8M images | 19 | 86M | PubMed + Twitter |
| Hibou L* | Nechaev 2024 | 1139k slides | 10 | 303M | Proprietary |
| Phikon | Filiot, 2023 | 6k slides | 13 | 86.4M | TCGA |
| Prov-GigaPath* | Xu et al., 2024 | 171k slides | 31 | 1011M | Proprietary |
| UNI* | Chen 2024 | 100k slides | 20 | 307M | Mass-100 |
| ConvNext | Liu 2022 | 1.24M images | N/A | 89M | ImageNet |
| DINOv2* | Oquab 2024 | 142M images | N/A | 86M | ImageNet + Web |
| EfficientNetV2S | Tan 2021 | 1.24M images | N/A | 22M | ImageNet |
| ResNet50 | He 2015 | 1.24M images | N/A | 25M | ImageNet |

### Beyond cancer – testing foundation models for non-neoplastic pathology

The pathologic basis of disease represents several diverse mechanisms.^14^ However, nearly all pathology benchmarks are cancer-based. Of the foundation models we included, 9/9 CONCH datasets, 12/12 GigaPath datasets, 10/10 Hibou datasets, 14/14 Phikon benchmarks and 26/27 UNI benchmarks were cancer related tasks, including cancer detection, typing, mutation prediction, segmentation, survival prediction, or tumor infiltrating lymphocyte related.^4,6–8,15^ The sole non-cancer benchmark was heart transplant rejection in UNI. Driven by their biological hallmarks, cancers share overlapping morphology including hypercellularity, frequent mitoses, elevated nuclear-cytoplasmic ratio, nuclear atypia, and necrosis.^16–20^ Thus the benchmarks, despite their number, examine a relatively small portion of the morphologic feature space in human pathology. Neoplasia is among the leading cause of death in the United States, but other leading causes like heart disease and stroke, chronic obstructive pulmonary disorder, Alzheimer disease, diabetes, Lupus nephritis, cirrhosis, and pneumonia reflect other types of pathologic processes.^21^ Benchmarks of ischemic, inflammatory, infectious, developmental, metabolic, and degenerative disease are needed.

### The placenta

The placenta is the unique organ that supports fetal development in utero. Placental functions include driving oxygen and nutrient delivery, removing toxic metabolites, blocking ascending and hematogenous infection, and managing maternal tolerance of the partially non-self fetus.^22,23^ The placenta is comprised of diverse tissues including multi-layered membranes, umbilical cord, chorionic plate, villous core, and the maternal-fetal interface at the decidua. Placental pathology is performed for a variety of indications, including suspected infection, growth restriction, hypertension or diabetes in pregnancy, preterm birth, adherent placenta, abruption, or stillbirth.^24^ On examination, these presenting concerns are linked to a wide set of diagnoses, which are driven by one or more modes of pathogenesis, whether infectious, vascular, ischemic, thrombotic, metabolic, developmental, or inflammatory.^25^ The variety of placental diagnoses, high frequency of diagnoses, and commonness of the placental specimen make it well-suited to support benchmarks in non-neoplastic pathologic processes.^26^ Significantly for the present work, placenta has not been used in previous benchmarking efforts. Furthermore, placenta is not well represented in the training data for these models. CONCH is the only model described to contain placental data (see Extended Data 1 in ^15^), where it makes up a small proportion. Benchmarking in placenta is therefore an opportunity to test the generalizability of pathology foundation models. Our group, and others, have developed machine learning models for placental diagnoses.^27–32^ In our recent work, we showed promising results for pathology foundation models in placental applications.^33,34^ However, each work focused on only 1 conventional and 1-2 pathology foundation models. Given the lack of sufficient benchmarks in non-neoplastic pathology and opportunities in placental pathology, we developed multiple benchmarks.

### We propose benchmarks in these areas of placental pathology

#### Gestational age

Oxygen and nutrient exchange in the placenta take place in the villi. These finger-like projections juxtapose fetal capillaries with maternal circulation. Over the course of gestation, a process of maturation occurs in which the number of villi increases dramatically while individual villi narrow, cellularity fluctuates and in the stromal and syncytiotrophoblast layers change.^35,36^ Alterations in maturation can be seen with hypertensive disorders of pregnancy, gestational diabetes, or other pathologies.^37–41^ In our task, models classify the gestational age of patches from placentas with normal maturation at 24-42 weeks gestational age.^30^ This is a different and more challenging problem than that faced by pathologists, who are given the gestational age and asked to determine whether the microscopic appearance matches the stated age. Even then, pathologists show a high degree of interobserver variability.^30,42^

#### Region classification

We have generated a dataset of 30,000 patches of normal placental tissue representing 6 major regions present in the placenta – amnion, decidua, fibrin, glass (background), stem villi including chorionic plate, and terminal villi (**Figure 3**). This task is relatively straightforward given that patches are of normal tissues in high abundance. We tested Gestational Age estimation and Region Classification using K-nearest neighbors because it is typically robust in zero-shot scenarios, requires no large-scale parameter training, and allows a direct evaluation of how well the pre-trained embeddings cluster.

#### Umbilical cord inflammation

Intrauterine inflammation and systematic inflammation in the fetus may manifest as neutrophil extravasation through the walls of the vein and arteries of the umbilical cord and into Wharton’s jelly. Umbilical vessel inflammation is staged as fetal inflammatory response, while inflammation in Wharton’s jelly is diagnosed as funisitis.^25^ Funisitis is associated with adverse neonatal outcomes including early onset sepsis, cerebral palsy, and death.^43–45^ We tested untrained computer based image retrieval (CBIR) in this dataset.

#### Maternal inflammatory response, histologic chorioamnionitis

During pregnancy, the uterus is lined by placental membranes, which form a barrier against bacteria that colonize the genitourinary and gastrointestinal tracts. Rupture of membranes allows bacteria and bacterial products to ascend into the uterine cavity. This manifests clinically as fever, elevated maternal white blood cell count, and tachycardia. In the placenta, pathologists identify infiltration of neutrophils into the membranes as maternal inflammatory response (MIR). MIR is frequent, being present in ~30% of placentas examined at our institution in 2024. We recently showed that cell classification and quantification of neutrophil infiltrates in the membranes can link histologic findings with systemic inflammation.^46^ In the present work, we extract patches with known numbers of neutrophils and train models to estimate the number. Identifying individual cells and classifying individual cells as neutrophils at high power are two relatively straightforward processes. However, estimating the number of neutrophils in a 20X field is more challenging.

#### Villous infarction, intervillous thrombus, perivillous fibrin deposition

Several macroscopic lesions can be identified within the placental disc, of which the most common are villous infarction, perivillous fibrin deposition (PVFD), and intervillous thrombi (IVT, **Figure 6**). Villous infarctions are areas of interrupted maternal circulation associated with hypertension in pregnancy, preeclampsia, growth restriction, stillbirth, and risk of cerebral palsy.^47–51^ Perivillous fibrin deposition (PVFD) may be focal, where it is thought to represent a reparative response to turbulent flow, or massive (MPVFD) where it can be associated with fetal growth restriction and stillbirth.^52–56^ Intervillous thrombi (IVT), are foci of clotted blood in the intervillous space, which are seen in ~10% of placentas over many contexts. We developed a model to distinguish these three lesions from one another and from sections of normal placental disc.^57^ We developed a dataset of 331 cases with normal parenchyma, 176 with villous infarction, 178 with intervillous thrombus, and 147 with perivillous fibrin deposition, with 1 slide per case.

## METHODS

The study was approved by the Institutional Review Board of Northwestern University Feinberg School of Medicine in Chicago, Illinois, as STU00214052.

### Gestational Age Zero Shot Regression

Annotated regions from Mobadersany et al., were used.^30^ H&E-stained slides of placental villi from 154 patients were scanned with a Hamamatsu Nanozoomer at 20x magnification. Image regions were hand-annotated, extracted at 10x, and color-normalized using Reinhard’s method.^58^ The dataset was split at the patient level 60:20:20 into training, validation and test sets, stratified by gestational age. All regions from the same patient were assigned to the same data set. Regions were tiled into non-overlapping 224×224 pixel patches. Features were extracted. KNN regressor was trained using scikit-learn.^59^ N-neighbors was determined using 10-fold cross validation within the training dataset. Metrics were performed on the test set and mean absolute error (MAE), the difference between estimated and actual gestational age, r^2^, and slope of the regression line between estimated and actual gestational age.

### Placenta Region Zero Shot Classification

For the Region Classification task, the overall pipeline is largely consistent with that of the Gestaltnet Zero Shot Regression task. The differences are as follows: Slides were from 207 placentas, stained with H&E and digitized at 40X using Leica GT450 scanners. The set was split 60:20:20 into training, validation, and test sets. Regions were manually annotated and extracted at 20X. Images were cropped to 128×128 pixel patches. Patches were stretched to 224×224 pixels, then input to feature embedding. Classification was performed using a KNN classifier. The primary metrics include patch-level balanced accuracy on the test set, and silhouette score of the embedding. t-stochastic neighbor embedding (t-SNE) plots were generated to visualize clustering.

### Umbilical Cord Inflammation CBIR

There were 11887 224×224 pixel patches at 20X from 598 slides.

Features were extracted. CBIR was performed using Euclidean distance, and retrieved patches could not be from the query slide. We measured recall and precision at k=1-8. Embeddings were compared using uniform manifold approximation and projection (UMAP)

### Neutrophil Infiltration Estimation

For the Neutrophil Infiltration task, the overall pipeline is largely consistent with that of the gestational age Zero Shot Regression task. The differences are as follows: Slides were digitized on a Leica GT450 and cell classifications was performed for 63 placental membrane slides from.^46^ The set was split into training, validation, and test sets with an 80:10:10. 224×224 pixel regions were extracted at 20X magnification and used as an input to the feature extractor. Neutrophil counts were estimated using a multilayer perceptron regressor with parameter search with hidden layers sizes of (256,32) or (128,32), stochastic gradient descent of Adam solver, initial learning rate of 1e-3, 5e-4, or 1e-4, learning rate that was constant, inverse scaled, or adaptive, and maximum iterations of 50, 100, or 200.^59^ 100 random parameter sets were drawn for each embedding. The model with the best performance on the validation set was used for the test set. The primary metrics include mean absolute error and correlation (r^2^) between estimated and actual number of neutrophils.

### Whole slide classification of macroscopic lesions

The dataset has been previously described.^31,32^ It consists of 832 whole slide images scanned on a Leica GT450 at 40X. The dataset was split 70:15:15 into training, validation, and testing. After tissue masking using Otsu’s method, features were extracted at 10X.^60^ We used an attention-based multiple instance learning approach.^61^ Hyperparameter search was performed using random search implemented by the Ray tune package.^62^ We used a broad set of hyperparameters for model design and optimization (**Supplementary Table 1**). For each foundation model, we performed 1000 trials, with minimum of 15 epochs and early stopping based on plateaued balanced accuracy in the validation set. Model performance was evaluated in the held-out test set. Performance was measured based on balanced accuracy of the best model and 50^th^ best model. The distribution of balanced accuracy for the top 100 model for each embedding were compared using Kruskal-Wallis test with post-hoc Mann-Whitney U tests, performed in scipy.^63^ 36 comparisons are made for a significance threshold of 0.00139.

## RESULTS

### Zero shot estimation of gestational age

Representative patches from 24-42 weeks are shown in **Figure 1**. Performance of individual models is shown in **Figure 2, Table 2**. UNI had the lowest MAE at 2.47 weeks, while none of the non-pathology models had MAE below three weeks. r^2^ was inversely correlated with MAE and was highest with UNI.

**Table 2.** Model performance across benchmarks.

|  | Gestational age |  | Region classification |  | Cord CBIR precision@8 |  | Neutrophil infiltration |  | Macroscopic lesions balanced accuracy |  |
| --- | --- | --- | --- | --- | --- | --- | --- | --- | --- | --- |
| Model | MAE | r<sup>2</sup> | Accuracy | Silhouette score | Overall | Funisitis | MAE | r<sup>2</sup> | Best | 50<sup>th</sup> best |
| CONCH | 3.01 | 0.561 | 0.829 | **0.182** | 0.813 | **0.341** | 1.43 | 0.653 | 0.888 | 0.726 |
| Hibou L | 2.60 | 0.640 | 0.894 | 0.159 | 0.782 | 0.243 | 1.27 | 0.736 | 0.907 | 0.788 |
| Phikon | 2.85 | 0.563 | 0.860 | 0.179 | 0.779 | 0.249 | 1.28 | 0.761 | **0.922** | 0.767 |
| Prov-GigaPath | 2.53 | 0.664 | 0.872 | 0.104 | 0.789 | 0.279 | 1.28 | 0.730 | 0.896 | 0.791 |
| UNI | **2.47** | **0.703** | **0.895** | 0.120 | **0.815** | 0.333 | **1.21** | **0.767** | 0.908 | **0.821** |
| ConvNext | 3.45 | 0.391 | 0.782 | 0.079 | 0.745 | 0.298 | 2.02 | 0.320 | 0.907 | 0.767 |
| DINOv2 | 3.44 | 0.372 | 0.762 | 0.120 | 0.756 | 0.332 | 1.98 | 0.349 | 0.895 | 0.755 |
| EfficientNet | 3.40 | 0.409 | 0.798 | 0.137 | 0.755 | 0.307 | 1.51 | 0.618 | 0.918 | 0.789 |
| ResNet | 3.69 | 0.306 | 0.723 | 0.084 | 0.714 | 0.208 | 2.71 | -0.124 | 0.666 | 0.542 |

**Figure 1.**
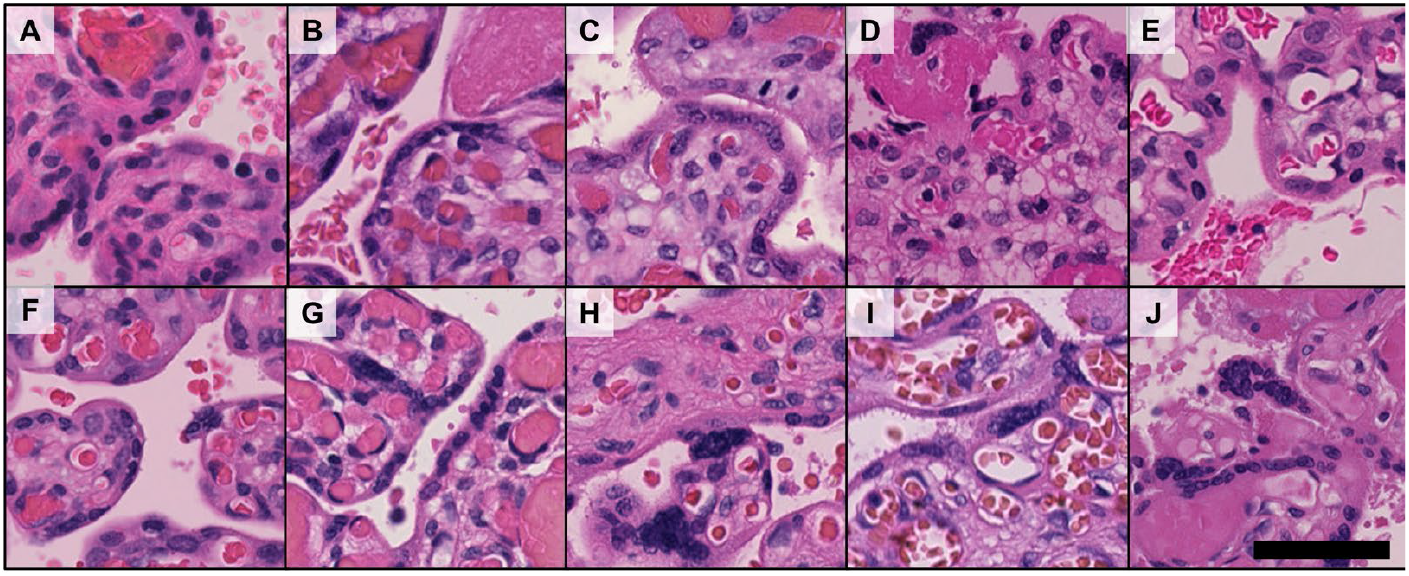
placental morphology across gestational age. Patches of placental villi from 24 weeks gestation (A) to 42 weeks gestation (J) show decreasing villous diameter, denser stroma, and increased syncytial knots with greater gestational age. Original magnification 10X, scale bar, 100 µm.

**Figure 2.**
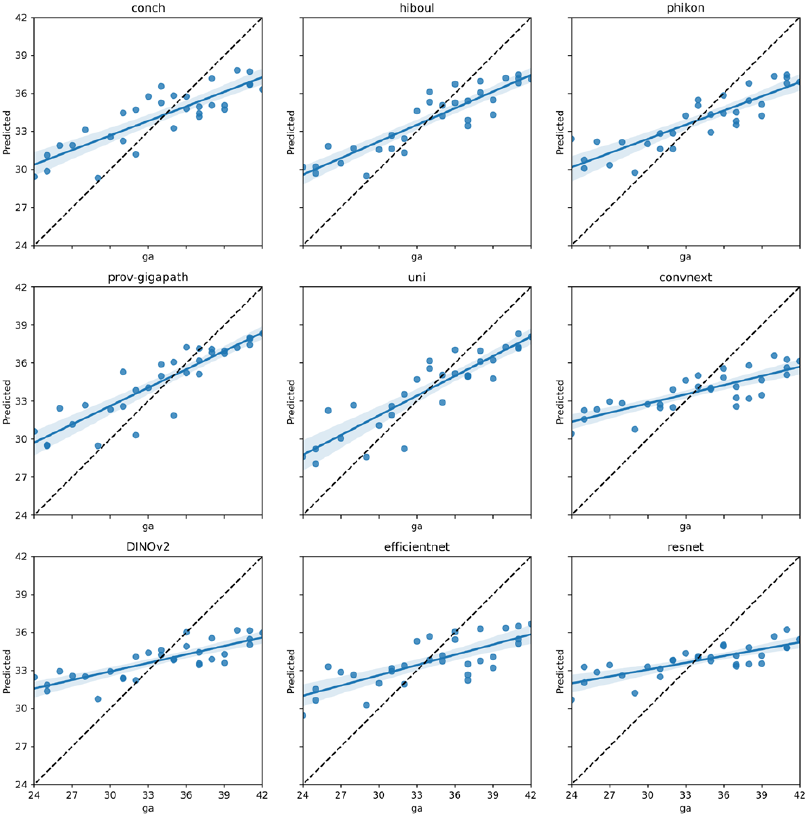
Zero Shot GA estimation. Scatter plots of patient-level predicted vs. actual gestational age are shown with regression lines. UNI showed the lowest MAE and highest r2. Dotted lines show ideal performace (actual = expected).

### Zero shot region classification

Representative patches from the six classes are shown in **Figure 3**. Performance of models is shown in **Table 2**. The pathology foundation models showed accuracies between 0.829 and 0.895, and every pathology model was superior to all non-pathology models. Silhouette scores for tSNE embeddings ranged from 0.079 for ConvNext to 0.182 for CONCH (**Figure 4**).

**Figure 3.**
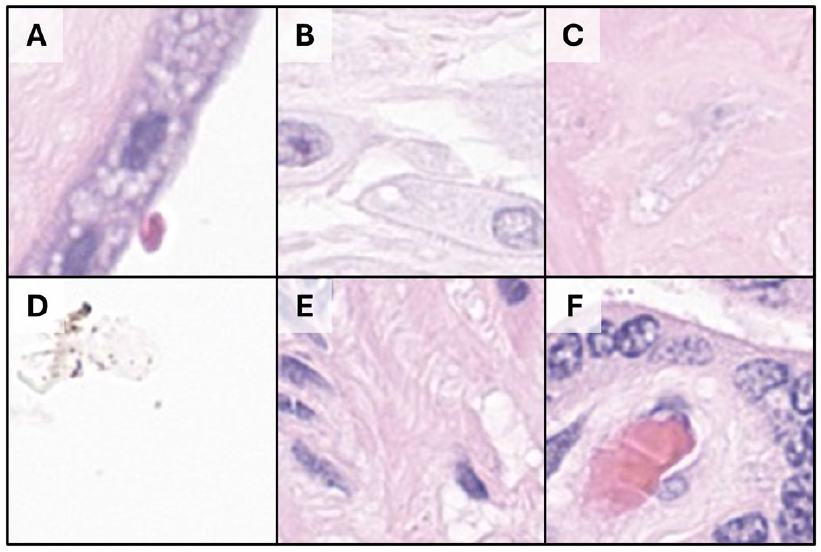
Placental disc region patches: Images of amnion (A), decidua (B), perivillous fibrin (C), glass with debris (D), stem villi (E) and terminal villi (F). Images are 128×128 pixels, original magnification 40X.

**Figure 4.**
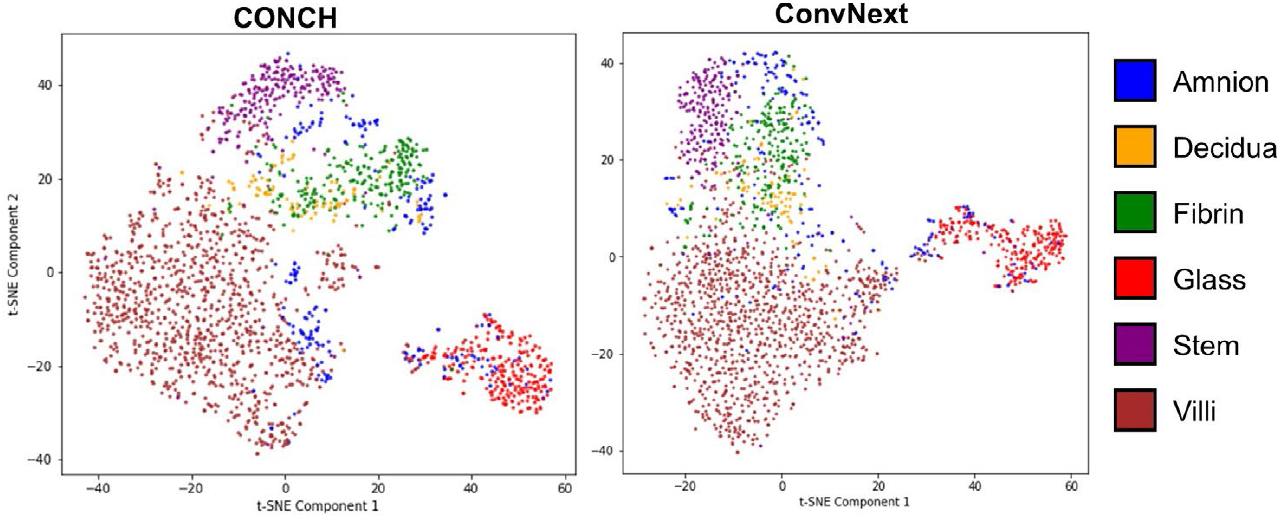
Placental region clustering: t-SNE plots of embedding generated by CONCH (left) and ConvNext (right), which have the best and worse sillhouete scores of 0.182 and 0.079, respecitvely. ConvNext intersperses amnion patches with other regions, while conch shows clearer separation of fibrin and decidua. Glass is readily separable by both models. (See Supplementary Figure 1 for tSNE plots from the other tested models).

### CBIR in umbilical cord

UNI had the best overall precision@8, with 0.815, and every pathology foundation model outperformed every non-pathology model (**Figure 5**). However, performance was heterogeneous across patch types, with particularly poor performance in the inflamed Wharton’s Jelly (funisitis) with the highest precision@8 of 0.341 for CONCH. The inflamed vessel wall classe also showed poor performance.

**Figure 5.**
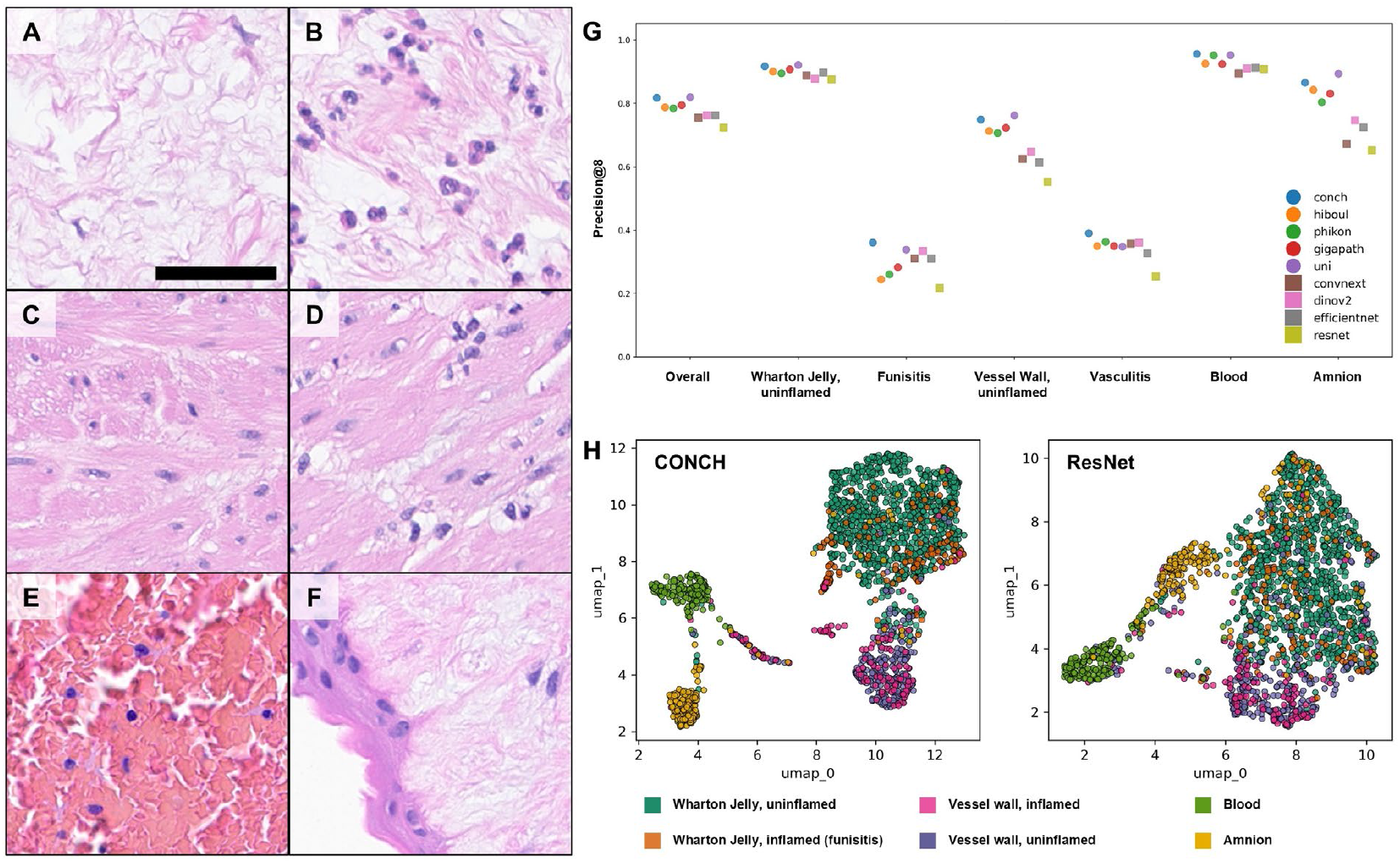
Umbilical cord content-based image retrieval: Sample images of Wharton’s Jelly without inflammation (A), inflamed Wharton’s Jelly, diagnostic of funisitis (B), uninflamed umbilical vessel wall (C), inflamed vessel wall, (D), blood (E), and umbilical amnion overlying Wharton jelly (F). Comparative precision at 8 overall and for each tissue class. Round markers indicate pathology foundation models, square markers are non-pathology models. UMAP plots show distinction between vessel wall, Wharton’s Jelly, blood and amnion, but inflamed and uninflamed patches are interspersed. (H). Scale bar = 50 um.

### Neutrophil infiltration estimation

Patches and regression predictions from UNI and ResNet are shown in **Figure 6**. UNI had the lowest MAE and highest r^2^. ResNet had the worst performance (**Table 2**). Like other tasks, every pathology foundation model outperformed every non-pathology model.

**Figure 6.**
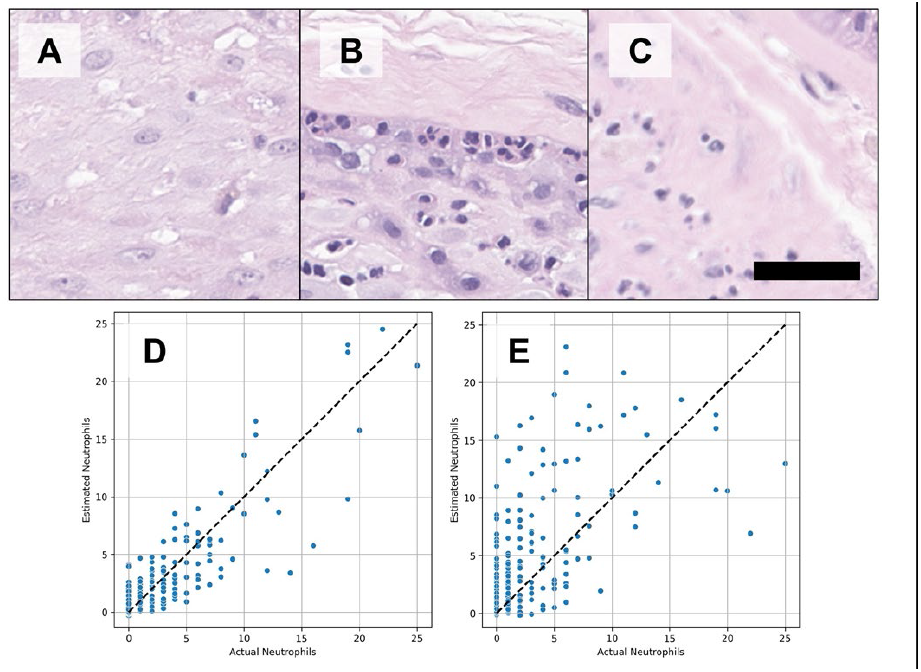
Neutrophil count estimation: Patches of placental membrane showing uninflamed decidua (A), neutrophils in the decidua aligning at the subchorion (B), and neutrophil infiltration into the fibrous chorion and amnion (C). Actual vs. estimated neutrophil counts for UNI (D) and ResNet (E), the best and worst performing models, respectively. Dotted line shows perfect estimation (actual = estimated). Original magnification 20X, scale bar 50 µm

### Whole slide classification of macroscopic placental lesions

Representative images are shown in **Figure 7**. We trained 1000 models using different hyperparameters. The single best model used Phikon embeddings, with a balanced accuracy of 0.922. Unlike other tasks, non-pathology models did well, with the top ConvNext, EfficientNet, and DINOv2 models each outperforming the top CONCH model, and the top EfficientNet model 2^nd^ best overall. High performance for a single model may be due to fortuitous model design and initialization. A model that produces more robust embeddings should do well with less-than-perfect design and initialization, so we examined the drop between the best and 50^th^ best model. For example, the drop off between the best Phikon model and 50^th^ best was 0.922 to 0.767, while that for UNI was only 0.908 to 0.821. It is arguable that a better model will show more consistency in performance. To examine this, we plotted and compared the performance of the top 100 models trained with each embedding (**Figure 7e**). In brief, while ResNet as clearly the worst, the top trio of UNI, Prov-GigaPath and Hibou did not statistically differ. Among the non-pathology models, EfficientNet and ConvNext were each not statistically different from one or more pathology foundation models. Full pairwise comparisons are given in **Supplementary Table 2**.

**Figure 7.**
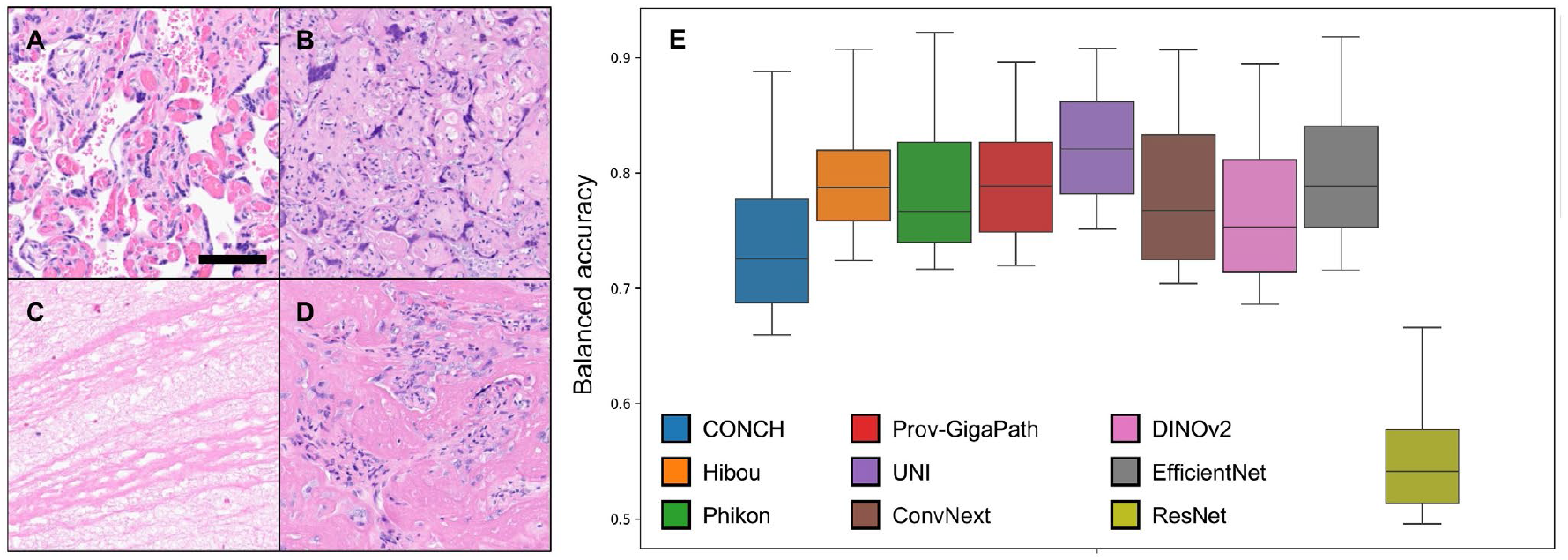
whole slide classification of macroscopic lesions: Models were trained to distinguish whole slides of normal placenta (A), villous infarction (B), intervillous thrombus (C), and perivillous fibrin deposition (D) using attention-based multiple instance learning. Boxplot of balanced accuracies for the top 100 models trained using each embedding (E). See main text and **Supplementary Table 2** for statistical comparisons. Original magnification 10X, scale bar: 100µm

### Analysis of model performance

We analyzed the performance of models based on their number of parameters, training set size, and inclusion of pathology content. The number of models is insufficient for rigorous statistical analysis, but some notable trends emerge. For both gestational age metrics and for region classification balanced accuracy, training on pathology images was the key differentiator (**Figure 8**). The trend was most evident seen for CONCH and Phikon, which consistently outperformed ConvNext and DINOv2, even though these models all have 86-89 million parameters. The effects of data set size and embedding dimension were more complicated (**Supplementary Figure 2**). Within the pathology models, there is a weak positive relationship between dataset size and performance. For neutrophil estimation, pathology models did better, though EfficientNet was competitive. For macroscopic lesions, ResNet50 performs the worst, but the other non-pathology models were competitive with the pathology models.

**Figure 8.**
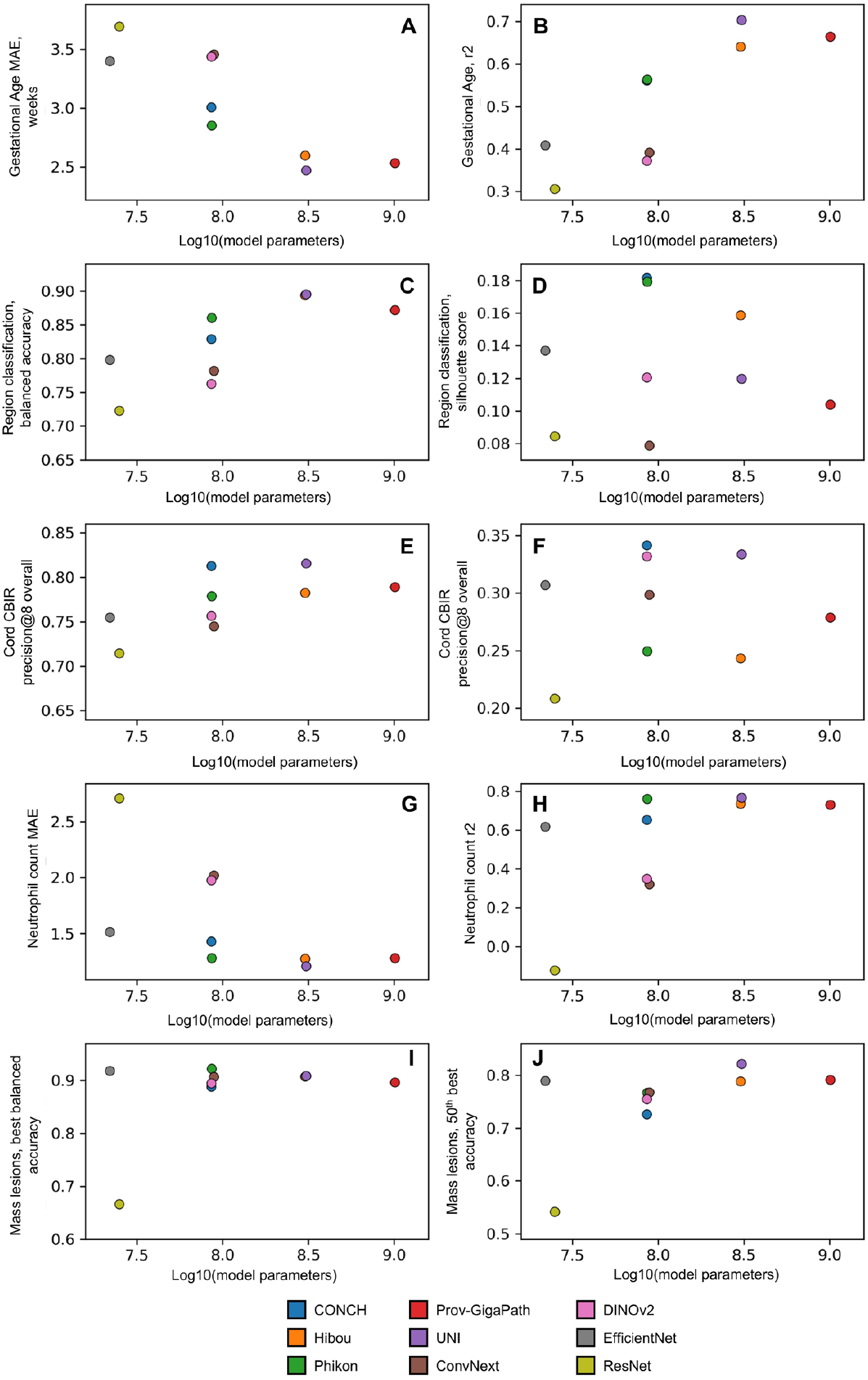
Analysis of model performance: Performance of models in gestatonal age (A & B), region classification (C & D), cord retrieval (E&F), neutrophil estimation (G & H) and mass lesion classification (I & J). For gestational age, region classification, and neutrophil count, there is a correlation between larger model size and higher performance, though with saturation. The relationship between model size and region sillhouette score, cord precision, and mass lesion classification is more complex.

## DISCUSSION

### Recitation

We tested several pathology foundation models and non-pathology model on benchmarks in placental pathology. We show that pathology-specific foundation models overall show better performance, but that performance is similar within the group. We show that the advantage is most pronounced for zero shot K-nearest neighbors estimation and is blunted when multilayer perceptrons or multiple instance learning paradigms are used.

### Gestational age

In zero shot estimation of gestational age, every pathology-specific model outperformed every non-pathology model, with UNI showing the best MAE at 2.47 weeks. This is worse than our previously published performance of 1.09 weeks in Mobadersany et al., using end-to-end training and 1.62 weeks in Goldstein et al. using multiple instance learning.^30,64^ However, 2.47 weeks is better than the 2.72 week MAE produced by human pathologists when they are asked to estimate gestational age – admittedly not a task for which they have been trained.^65^

### Region classificatio

In a zero-shot region classification task, every pathology model outperformed every non-pathology model. There were similar patterns to gestational age estimation where larger model size and training set size were associated with higher performance. We used silhouette score as a measure of clustering performance, on the theory that stronger clustering is indicative of better alignment in feature space. This approach has been used in unsupervised learning, where the label of individual patches is unknown.^66^ Interpretation of these results may be challenging. Silhouette score relies upon Euclidean distance – models with a higher embedding dimension may inherently have result in larger distances.

### Umbilical cord CBIR

Overall retrieval performance was quite good, given the stringency of precision@8. However, performance for inflamed Wharton’s Jelly or vessel wall was quite poor. We used an untrained approach to CBIR. Unlike zero shot classification, CBIR has a strong role in creating explanations, a role which is likely best served by the natural growth of an archive of previously diagnosed cases.^67–70^

### Neutrophil estimation

Similarly to gestational age and region classification, pathology foundation models performed best on neutrophil estimation, although the relationship with model size and number of patches was less strong (Figure 8E and F).

### Mass lesion classification

Perhaps due to the complexity of the multiple instance learning design, the advantage of pathology foundation models was blunted. Our previous best performance for this task was a balanced accuracy of 0.904, achieved using the EfficientNet feature extractor and manual tuning of the hyperparameters.^32^ Several of our models outperformed this benchmark, including EfficientNet – this highlights the value of random search and suggests out search was sufficiently exhaustive for meaningful analysis.

### Does pathology training matter?

ResNet50 was released in 2015, contains far fewer parameters than pathology foundation models, was trained on a smaller dataset and uses CNN rather than ViT architecture. Yet, Phikon and UNI tested against ResNet in their initial publications, while CONCH, Hibou-L, and Prov-GigaPath did not test against any non-pathology models.^4,6–8,15^ Of three recent benchmarking papers, one tested against ResNet and the others included no non-pathology models.^13,71,72^ This likely overstates the performance advantage of pathology-specific foundation models over non-pathology models. Our approach is different. Beyond ResNet, we chose to include EfficientNet, and ConvNext in our study because we used those models in recent studies to good effect. We included DINOv2 because its training protocol underlies several of the leading foundation models. The gap between these models and that pathology foundation models is much smaller.

Our findings with whole-slide lesion diagnosis are telling – with the more complicated model design built atop foundational embeddings, non-pathology models provide sufficiently valuable information. Content-based image retrieval seems to buck this trend – pathology foundation models and non-pathology models alike were challenged by inflamed tissues, which consistently overlapped their uninflamed counterparts in feature space. We cannot help but speculate that the (near-)exclusive focus on neoplastic pathology explains this finding.

### Does composition matter?

Among pathology foundation models tested on existing benchmarks, reported performance is comparable with small differences, particularly among the models with the largest datasets and models.^12,13,73^ Of observable differences, dataset composition plays a role. For example, Filiot et al. found that a colon adenocarcinoma-specific model, trained on 4.4 million patches outperformed Prov-GigaPath on colon cancer tasks, even though Prov-GigaPath contains 277 million patches of bowel.^4,73^ Similarly, foundation models with a higher *proportion* of lung slides in their training data do better at lung-specific benchmarks.^13^

Placental pathology makes an interesting contrast to this trend, since most pathology foundation models are not documented to include placenta. We included CONCH in our study because it is the only pathology foundation model documented to include placenta images.^15^ We estimate that ~30,000, or ~1.67% of the of the 1.8 million images in CONCH are of placenta. It is difficult to evaluate the impact of the placenta images on CONCH’s performance. CONCH is among the smaller pathology foundation models, with fewer parameters and a smaller dataset size. The only conclusion we can draw is that, insofar as the placenta images are improving CONCH, they are insufficient to raise it above larger pathology foundation models that do not include placenta. This suggests that, for organs that are poorly represented in pathology foundation training data, selection of a model that includes a small amount of the target organ is a secondary consideration.

## CONCLUSION

In conclusion, we tested pathology and non-pathology foundation models on a set of placental benchmarks. In line with previous studies, we found that pathology foundation models were generally better with small differences based on the size of model and training data. In inflammatory tasks, trainable patch classification, and whole slide diagnostic studies, the advantage of pathology models was blunted. Future work could examine the impact of incorporating more placental training data.

## DATA AVAILABILITY STATEMENT

Benchmarking data are available from Northwestern after execution of a data use agreement.

## FUNDING AND CONFLICT OF INTEREST

Goldstein is supported by National Institutes of Health (NIH) R01EB030130 and UG3OD035546. Cooper is supported by NIH U24NS133949, R01LM013523, and U01CA220401. Infrastructure used by this work, including REDCap, is supported by UL1TR001422.

Cooper also reports honoraria from Risk Appraisal Forum, Lynn Sage Breast Cancer Foundation, Jayne Koskinas Ted Giovanis Foundation for Health & Policy and Consultation for Tempus. The other authors have no relevant financial interest in the products or companies described in this article.

## Notes

### Competing Interest Statement

Cooper reports honoraria from Risk Appraisal Forum, Lynn Sage Breast Cancer Foundation, Jayne Koskinas Ted Giovanis Foundation for Health & Policy and Consultation for Tempus. The other authors have no relevant financial interest in the products or companies described in this article.

