## Supplemental Table 1 for "Benchmarking pathology foundation models for non-neoplastic pathology in the placenta"

**SUPPLEMENTARY TABLE 1, PARAMETER SEARCH FOR CLASSIFICATION OF MACROSCOPIC LESIONS**

| **Hyperparameter** | **Options** |
| --- | --- |
| Backbone layers | [1,2] |
| Attention layers | Fixed at 1 |
| Task layers | Fixed at 1 |
| Layer size | [512, 256, 128, 64]* |
| Layer activation | [‘elu’, ‘linear’]** |
| Dropout | [0, 0, 0, 0.1, 0.25]*** |
| L1 norm | [0, 1e-3, 1e-2] |
| L2 norm | [0, 1e-3, 1e-2] |
| *: Layer size, activation, and dropout are randomly selected per layer. **: The final backbone layer is fixed as (sigmoid, tanh) for gated attention. There is a final task layer with 4 neurons and softmax activation. ***: 3/5 layers have no dropout, 1/5 have dropoout = 0.1, 1/5 have dropout = 0.25 | |
