## Supplemental Table 2 for "Benchmarking pathology foundation models for non-neoplastic pathology in the placenta"

Performance of models in the multiple instance learning task:

| model | best_acc | 5th_best_acc | 10th_best_acc | 25th_best_acc | 50th_best_acc |
| --- | --- | --- | --- | --- | --- |
| conch | 0.888 | 0.857 | 0.828 | 0.778 | 0.726 |
| hibou | 0.907 | 0.881 | 0.865 | 0.822 | 0.788 |
| phikon | 0.922 | 0.877 | 0.851 | 0.827 | 0.767 |
| prov-gigapath | 0.896 | 0.868 | 0.858 | 0.829 | 0.791 |
| uni | 0.908 | 0.891 | 0.881 | 0.862 | 0.821 |
| convnext | 0.907 | 0.877 | 0.864 | 0.833 | 0.767 |
| dinov2 | 0.895 | 0.875 | 0.855 | 0.816 | 0.755 |
| efficientnetv2s | 0.918 | 0.876 | 0.867 | 0.844 | 0.789 |
| resnet50 | 0.666 | 0.652 | 0.613 | 0.578 | 0.542 |

Pairwise P-values in a Mann-Whitney U test. P-values are unadjusted for multiple comparisons

| model | vs_conch | vs_hibou | vs_phikon | vs_prov-gigapath | vs_uni | vs_convnext | vs_dinov2 | vs_efficientnetv2s | vs_resnet50 |
| --- | --- | --- | --- | --- | --- | --- | --- | --- | --- |
| conch |  | 2E-11 | 2E-08 | 2E-10 | 2E-18 | 1E-06 | 7E-04 | 1E-11 | 3E-34 |
| hibou | 2E-11 |  | 1E-01 | 7E-01 | 6E-05 | 2E-02 | 2E-04 | 7E-01 | 3E-34 |
| phikon | 2E-08 | 1E-01 |  | 3E-01 | 5E-07 | 3E-01 | 5E-03 | 6E-02 | 3E-34 |
| prov-gigapath | 2E-10 | 7E-01 | 3E-01 |  | 2E-05 | 5E-02 | 6E-04 | 4E-01 | 3E-34 |
| uni | 2E-18 | 6E-05 | 5E-07 | 2E-05 |  | 1E-07 | 3E-10 | 6E-04 | 3E-34 |
| convnext | 1E-06 | 2E-02 | 3E-01 | 5E-02 | 1E-07 |  | 6E-02 | 8E-03 | 3E-34 |
| dinov2 | 7E-04 | 2E-04 | 5E-03 | 6E-04 | 3E-10 | 6E-02 |  | 5E-05 | 3E-34 |
| efficientnetv2s | 1E-11 | 7E-01 | 6E-02 | 4E-01 | 6E-04 | 8E-03 | 5E-05 |  | 3E-34 |
| resnet50 | 3E-34 | 3E-34 | 3E-34 | 3E-34 | 3E-34 | 3E-34 | 3E-34 | 3E-34 |  |
