## Supplemental Figure 1 for "Benchmarking pathology foundation models for non-neoplastic pathology in the placenta"

### Slide 1
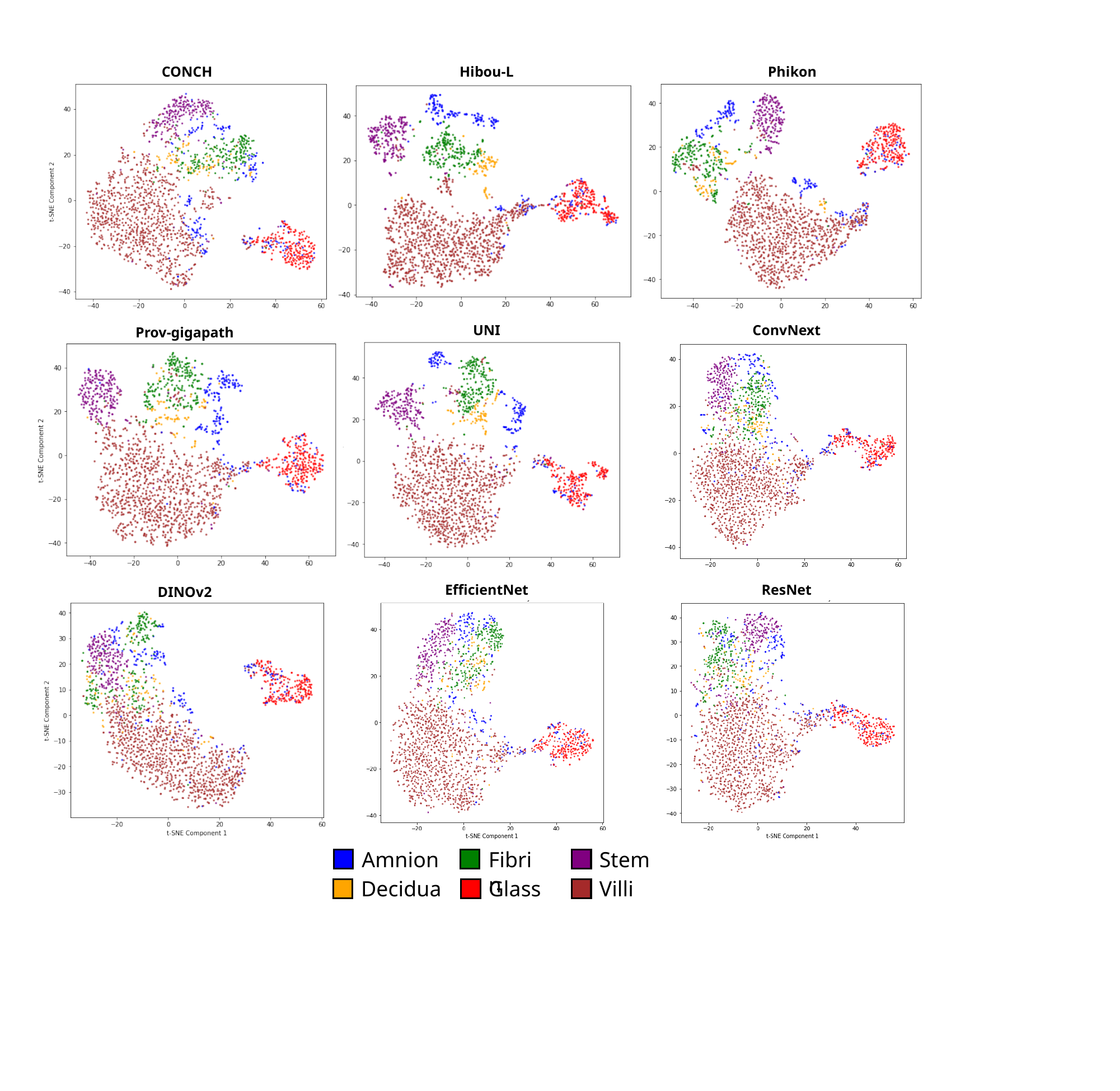

CONCH
Hibou-L
Phikon
ConvNext
UNI
Prov-gigapath
ResNet
EfficientNet
DINOv2
Amnion
Fibrin
Stem
Decidua
Glass
Villi
